# The effect of the fortifying the spleen, clearing heat, activating blood method on chronic atrophic gastritis: a real-world study

**DOI:** 10.1101/2019.12.25.19015800

**Authors:** Zhihua Zheng, Yi Wen, Yanhua Yan, Zhiheng Xu, Kechao Nie, Xu Chen, Fengbin Liu, Jinglin Pan, Peiwu Li

## Abstract

**Introduction:** Chronic atrophic gastritis (CAG) is a precancerous disease that is difficult to treat. Even after eradication of the *Helicobacter pylori* (HP) infection, complete resolution of the symptoms is difficult to achieve. The fortifying the spleen, clearing heat, activating blood method (FSCHABM) has an excellent curative effect in the treatment of CAG. A real-world study is particularly suitable for researching the treatment of CAG, but there are currently no reports on CAG trials. Our aim is to design a high-quality trial to investigate the efficacy and safety of FSCHABM in treating CAG patients.

**Methods and analysis:** This protocol is designed as a real-world study for 10 years. A total of 5000 participants will be assigned to a FSCHABM treatment group or a non-FSCHABM treatment group at a 1:1 ratio at the first Affiliated Hospital of Guangzhou University of Chinese Medicine. Patients are given 1-2 courses of a 24-week-long treatment. The participants will be followed up for observation and measurement of the following indicators: the primary outcome is the histopathological indicator; the secondary outcome includes evaluation of gastric lesions, syndrome curative effect, evaluation of symptoms, evaluation of quality of life, evaluation of anxiety and depression, economic evaluation and other indicators. This is the first real-world study evaluating the therapeutic effect of FSCHABM in the treatment of CAG in clinical practice. This protocol can provide a reference for future multi-center, randomized, controlled trials.

**Strengths and limitations of this study:** it is the first time to carry out TCM study related to CAG by using the real-world study. A large number of patients and a long-term study can effectively reflect the effect of TCM treatment and reduce the bias. The precise research protocol makes the whole research more accurate and reliable. The limitations of implementing this protocol are expending a lot of time, manpower and economic resources inevitably.

**Ethics and dissemination:** this study was approved by Ethical committee of the first Affiliated Hospital of Guangzhou University of Chinese Medicine. Study findings will be shared with participants, healthcare providers, and policymakers through research reports, conference presentations, and the Internet. The results will also be disseminated through publication in peer reviewed journals.

**Trial registration:** The registration number, ChiCTR1900027177, was assigned by the Chinese Clinical Trial Registry on 3 November 2019.

**Funding:** This work was supported by Guangdong natural science fund project, China (2019),No.2019A1515011145; Major research project of Guangzhou University of Chinese medicine, China (2019), No.A1-2606-19-110-007; The first affiliated hospital of Guangzhou University of Chinese medicine “innovation foster hospital” clinical research project, China (2019), No.2019IIT19; Guangdong natural science foundation (PhD) project, China (2017), No.2017A030310121; The first affiliated hospital of Guangzhou University of Chinese medicine “innovation foster hospital” innovation research team project, China (2017), No.2017TD05; The first affiliated hospital of Guangzhou University of Chinese medicine “innovation foster hospital” Youth scientific research talent training program, China (2015), No.2015QN09.

## Introduction

Chronic atrophic gastritis (CAG) is a common type of precancerous lesion, in which the repeated damage to the gastric mucosa epithelium causes a reduction of the intrinsic glands. Helicobacter pylori (HP) infection is the most important cause of CAG^[1,2]^, which is epidemiologically associated with the occurrence of gastric cancer (GC)^[3,4]^ and an unhealthy lifestyle, including alcohol abuse, tobacco abuse, high intake of sodium, preserved and spicy food^[5-7]^. A national multicenter cross-sectional study^[8]^ showed that 17.7% of the examined patients (1,573/8,892) was diagnosed with CAG at endoscopy. The annual incidence of GC for patients with CAG within 5 years of diagnosis was 0.1% ^[9]^. A global cancer investigation^[10]^ reported approximately 18.1 million new cancer cases and 9.6 million cancer deaths globally in 2018, with GC being the fifth most common cancer and the third cause of cancer death.

There are various treatments for CAG, including the eradication of HP, antacids, acupuncture, prokinetics and mucosal-protective agents^[8,11]^. For HP positive patients, eradication therapy is still the most basic and effective treatment for CAG^[12]^ that strongly improves the quality of life^[13]^, and promotes the revitalization of the gastric mucosa^[14]^. A follow-up study of 7.5 years indicated that eradication of HP in patients without precancerous lesions could significantly reduce the risk of gastric cancer^[15]^. Compared to chemical drugs, traditional Chinese medicine (TCM) has a unique advantage in the treatment of CAG and produces no serious side effects^[16]^. For example, Weiqi Decoction attenuated CAG with precancerous lesions by regulating the disturbed gastric mucosal blood flow and the HIF-1 signaling pathway^[17]^. Tian G et al^[18]^ adopted immune repertoire sequencing techniques to evaluate the effect of Modified Sijunzi Decoction for treating CAG. Results indicated that MSD could availably relieve CAG symptoms and improve pathological changes in CAG. According to the Consensus on TCM Diagnosis and Treatment of Chronic Gastritis (2017)^[19]^, the main pathogenesis of CAG is the spleen-stomach vacuity, qi stagnation and blood stasis, therefore, the fortifying the spleen, clearing heat, activating blood method (FSCHABM) is recommended for the entire treatment process of CAG.

TCM attaches importance to the concept of holism, and the TCM practitioner uses different treatment methods according to the constitution of the patient, the season and the area, providing a personalized treatment. Due to the limitations of randomized controlled trials (RCTs), this study design is unable to show the situation of each individual patient during the study of TCM treatments. A real-world study pays attention to the clinical application, especially the patient’s symptoms after treatment^[20]^. He Wei^[21]^ thinks the advantages and characteristics of TCM’s comprehensive therapy can be fully implemented under real world conditions. In addition, the evidence-based level of TCM treatment of CAG is low, and RCTs fails to represent the reality of TCM treatment. Thus, there is a need for a new clinical research method to provide high-level results-based evidence. Compared to RCTs, a real-world study is more suitable for clinical research of TCM.

## Methods and analysis

This current study is designed as a real-world study for 10 years which complies with the Declaration of Helsinki, Good Clinical Practice (GCP) Guidelines^[22]^ and the requirements of clinical trials by the Drug Administration Law of the People’s Republic of China, and will strictly abide by all laws governing TCM drugs. It is a real-world study that requires mass of data. Therefore, we determined 5000 sample as our research objects. A total of 5000 patients will be recruited voluntarily at the out- and in-patient department of the first Affiliated Hospital of Guangzhou University of Chinese Medicine. Depending on the type of medication the patients want to take without doctor’s intervention, the patients will be divided into FSCHABM group whose patients will receive FSCHABM treatment, and non-FSCHABM group whose patients receive western medicine, traditional Chinese medicine decoction (non-FSCHABM), acupuncture and other treatment methods. All patients will be requested to sign a written informed consent. The researchers participating in this experimental study will be trained strictly and individually to ensure the authenticity and accuracy of the research. This study is blind to outcome evaluators and data analysts. They are concealed the treatment plan of each subject and rejected to participate in the drug intervention process, in order to independently evaluated and analyzed the outcome of the subjects and their clinical data. Figure 1 shows the flow chart of this trial.

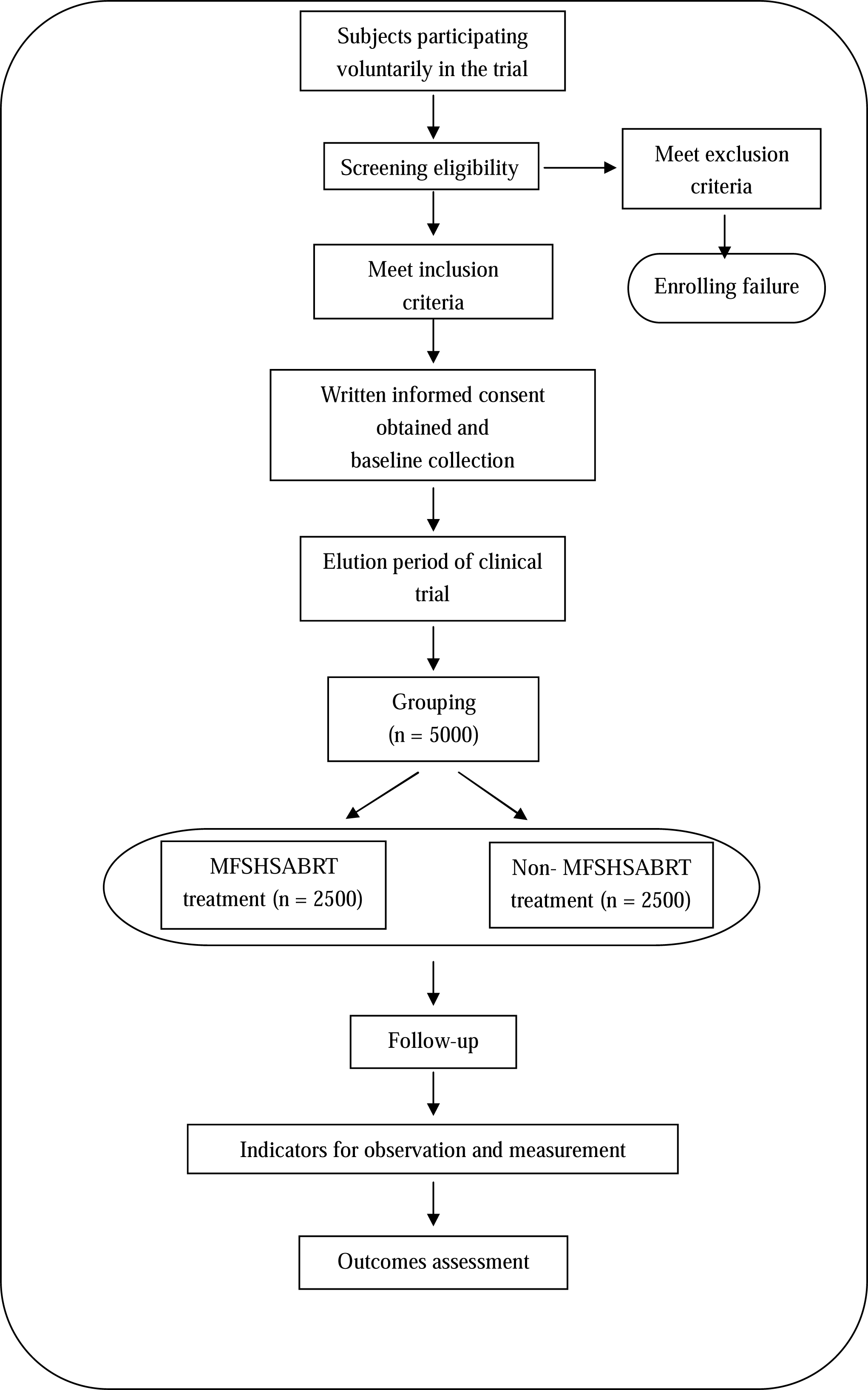

### Diagnosis in Western medicine

Referred to Chinese consensus on chronic gastritis (2017, Shanghai)^[11]^, CAG will be diagnosed by digestive endoscopy and pathological biopsy of gastric mucosa.

### Clinical manifestations

Most patients with CAG have silent clinical symptoms. These include non-specific gastrointestinal symptoms, such as upper abdominal discomfort, loss of appetite, belching, which can be accompanied by systemic or mental symptoms such as fatigue, wasting, forgetfulness, anxiety, etc. It has been reported that heartburn, regurgitation and early satiety are present in approximately 24%, 12%, and 10.1% of patients respectively ^[23]^.

### Endoscopic characteristics

Under the endoscope, the gastric mucosa can be seen between the mucous membrane as a red and white phase, white-based, wrinkled flattening or even nonexistent. Part of the mucous membrane blood vessels are observed, and can be accompanied by mucosa particles, nodules, and other manifestations.

Changes in mucosal inflammation seen by the gastroscopic doctor or special imaging methods under the endoscope should be combined with the results of pathological biopsy for the final judgment. Special imaging methods, such as magnifying endoscopy combined with staining and confocal laser microendoscopy, provide certain value in the diagnosis of CAG.

### Histopathological diagnosis

If the pathological biopsy of chronic gastritis shows inherent atrophy of the glandular body, CAG can be diagnosed, regardless of the number and degree of atrophy of the biopsy specimens. Clinicians can judge the scope and extent of gastric mucosa atrophy according to the pathological results combined with the gastroscopy.

### TCM syndrome diagnostic criteria of CAG

In acknowledgement of the consensus on diagnosis and treatment of CAG with TCM^[24]^, the TCM syndrome diagnostic criteria of CAG patients are listed as follows:

1. Pattern of liver and stomach qi stagnation: Main symptoms: distention or pain in the stomach; rib-side distention and pain. Secondary symptoms: symptoms induced or aggravated by emotional factors; frequent belching; choking sensation in the chest; thin white tongue coating; string pulse.
2. Pattern of intense liver-stomach fire: Main symptoms: clamoring stomach or burning pain in the stomach; string or rapid pulse. Secondary symptoms: heart vexation and irritable; clamoring and acid reflux in the stomach; bitter mouth and dry throat; dry stool; red tongue with yellow moss.
3. Pattern of weak spleen and stomach (pattern of cold spleen and stomach deficiency): Main symptoms: distention or dull pain in the stomach; the stomach likes to be pressed or warmed. Secondary symptoms: decreased food intake; dilute stool; fatigue and lack of strength; shortness of breath and laziness of speech; distended stomach after eating; pale tongue and weak pulse.
4. Pattern of spleen-stomach damp-heat: Main symptoms: stomach distension or pain; red tongue with yellow thick or greasy coating. Secondary symptoms: bitter mouth and halitosis; nausea or vomiting; heartburn; Sticky or dilute stool; slip and rapid pulse.
5. Pattern of insufficiency of stomach yin: Main symptoms: stomach oppression or burning pain; red tongue with less fluid and less coating. Secondary symptoms: loss of appetite and clamoring stomach; dry mouth; dry stool; thin body and decreased food intake; fine pulse.
6. Pattern of blood stasis in stomach collaterals: Main symptoms: fullness and compression in the stomach or pain in a specific location; the tongue is dark red or has petechiae or ecchymosis. Secondary symptoms: persistent stomachache; black stool; dark complexion; string and rough pulse.

Syndrome determination: the main symptoms must be present, and more than 2 secondary diseases be added to the diagnosis. In addition, the above syndromes may appear individually or simultaneously. In case of non-listed syndromes or simultaneous syndromes, syndrome differentiation can be standardized on the basis of the National Standard of the People’s Republic of China “Terminology of Clinical Diagnosis and Treatment of Traditional Chinese Medicine-Syndrome Part.” With the passage of time, the syndrome may change dynamically, requiring careful follow-up.

HP diagnosis and detection:

1. Invasive detection: rapid urea test; silver staining of biopsy pathological tissues.
2. Non-invasive detection: ^13^C urea breath test; ^14^C urea breath test.

Note: PPI, H2RA, bismuth, antibiotics and other drugs that may affect the results of the examination were discontinued for two weeks before the examination.

### Inclusion criteria

1. Age of 18-65 years old, regardless of gender.
2. CAG diagnostic by gastroscopy and histopathological examination.
3. Clear TCM syndrome types according to standard syndrome differentiation.
4. A one-minute rapid urease test, a ^13^C breath test or a ^14^C breath test were performed within the past month.
5. Voluntary provision of written informed consent.

### Exclusion criteria

1. Participated in other drug treatment studies within the past month.
2. Patients with autoimmune gastritis, peptic ulcer (A1-H2 phase), reflux esophagitis, gastric polyp, Gastric mucosa high-level intraepithelial neoplasia, gastrointestinal tumors, a history of gastrointestinal cancer and other digestive system disease.
3. Patients with serious organic diseases, such as: heart (NYHIII-IV grade of cardiac function, etc.), liver (decompensation stage of liver cirrhosis, etc.), kidney (uremia stage of chronic renal failure, etc.), lung (tuberculosis, acute attack of asthma, pulmonary infection, etc.), autoimmune system (active stage of systemic lupus erythematosus), etc.
4. Allergic constitution or allergy to multiple drugs.
5. Patients with severe mental diseases who cannot cooperate with researchers, such as schizophrenia, depression, dementia, etc.
6. Pregnant and nursing women.
7. Patients who do not cooperate.

### Withdrawal criteria

1. Loss of follow-up: ≥ 2 methods of contact (such as telephone, text message, mail) were followed up continuously for ≥ 3 times without reply.
2. Subjects voluntarily withdrew their informed consent and withdrew from the study.

### Rejection criteria

1. Misdiagnosis of the disease.
2. The proportion of missing items in the case report questionnaire exceeds 20%.
3. During the study period, intervention measures other than the scheme were combined.
4. Patients who fail to take the medicine according to instructions, for example, adjust the type, dosage, and usage of the medicine.

### Termination criteria

1. During the study period, the subjects developed new diseases that belong to the exclusion criteria.
2. Researchers found serious safety problems.
3. Research funds were insufficient to complete the study.
4. The administrative departments canceled the test.

### Therapeutic regimen

The therapeutic scheme of FSCHABM was formulated according to the Report Specification for Clinical Randomized Controlled Trials of Traditional Chinese Medicine Compound 2017. Patients with CAG were divided into sub-groups according to the main symptoms: asymptomatic patients; patients with stomachache; patients with stomach distension.

Core prescription: *Atractylodes Rhizome* 10 g, *Radix Pseudostellariae* 15 g, Poria 20 g, *Fructus Aurantii* 10 g, Perilla Stem 15 g, *Pericarpium Arecae* 15 g, *Herba Scutellariae Barbatae* 20 g, and *Curcumae Rhizoma* 10 g.

1. Asymptomatic (spleen and stomach weakness syndrome): core prescription.
2. Stomachache: core prescription + *Radix Aucklandiae* 10 g; Chicken bone broth 10 g.
3. Stomach distension: core prescription + Haematitum 15-30 g; *Lignum Aquilariae Resinatum* 3 g.

The dosage form of the above treatment scheme is a TCM decoction, which is prepared with 250 ml-300 ml of water twice a day for oral administration.

### Pattern identification and treatment administration

Pattern of liver and stomach qi stagnation: core prescription + Haematitum 15 g-30 g; *Lignum Aquilariae Resinatum* 3 g; *Dalbergia odorifera* 10 g.

Pattern of intense liver-stomach fire: core prescription + *Coptis chinensis* 5 g; *Evodia rutaecarpa* 3 g.

Pattern of spleen and stomach deficiency cold: core prescription + prepared aconite root 5 g-10 g; galangal root 10 g; pickled ginger 10 g.

Pattern of spleen-stomach damp-heat:

1. Pattern of equal severity of dampness and heat: the core prescription + *Poria cocos* is changed to *Smilax glabra* 30 g; talc 10 g to 20 g; calamus 20 g; *Agastache rugosa* 10 g; *Scutellaria baicalensis* 10 g.
2. Pattern of dampness predominating over heat: core prescription + *Herba Artemisiae Scopariae* 30 g; *Agastache rugosa* 10 g; slag leaf 15 g

Pattern of insufficiency of stomach yin: core prescription + *Ophiopogon japonicus* 15 g; *Dendrobium* 20 g; lily 30 g.

Pattern of blood stasis in stomach collaterals: core prescription + *Pollen Typhae* 5 g; *Oletum Trogopterori* 5 g; ground beetle 10 g.

### Differentiation of Symptoms

Intestinal metaplasia: core prescription + *Rhaponticum uniflorum* 20 g; *Sarcandra glabra* 20 g-30 g.

Intraepithelial neoplasia: core prescription + ground beetle 10 g; *Aspongopus* 10 g.

Accompanied by acid regurgitation: core prescription + *Coptis chinensis* 5 g; *Evodia rutaecarpa* 3 g.

Accompanied by belching: core prescription + haematite 15 g-30 g; *Lignum Aquilariae Resinatum* 3 g; *Dalbergia odorifera* 10 g.

Accompanied by poor appetite: core prescription + burnt hawthorn 10-20 g; medicated leaven 15 g-20 g.

Accompanied by throat blockage or sputum in the pharynx: core prescription + *Rhizoma Pinelliae Preparata* 10 g; snakegourd peel 20 g; *Allium macrostemon* 20 g.

Accompanied by chest tightness and discomfort: core prescription + sandalwood 10 g; snakegourd peel 20 g; *Allium macrostemon* 20 g.

Accompanied by loose stool: core prescription + white *Atractylodes* rhizome changed to fried *Atractylodes* 15 g; fried white lentils 30 g; *Agastache rugosa* 15 g.

Therapeutic regimen for Control Group

Western medicine and other TCM are used as control drugs, and the treatment plan is implemented according to Chinese Consensus on Chronic Gastritis (2017, Shanghai)^[25]^.

### Course of treatment

24 weeks of treatment is considered a course of treatment, and patients are given 1-2 courses of treatment.

### Drug administration

All drugs are clinical routine drugs provided and managed by the first Affiliated Hospital of Guangzhou University of Chinese Medicine. The western medicines are quasi-Chinese medicines. The pharmacy department of the center will implement standardized management of the drugs involved in the research to ensure the effectiveness and safety of each drug.

### Concomitant medication

Subjects should stop taking drugs for the treatment of CAG 1 month before entering this study. Subjects should take drugs according to the doctor’s treatment plan during the study period, and are forbidden to adjust the treatment plan by themselves. During the study period, the subjects can take drugs (e.g. antihypertensive drugs, hypoglycemic drugs, lipid-lowering drugs, etc.) to treat complicated diseases, which is recorded in detail in the medication records.

### Observation indicators

#### Epidemiological Indicators

1. Demographic data: subject’s name, age, sex, date of birth, occupation, place of birth, address, and contact information.
2. Disease-related factors: 1. drinking history, smoking history, working environment, intensity and time; 2. dietary preferences; 3. dietary habits; 4. use of NSAID drugs; 5. HP infection; 6. duration of digestive system symptoms; 7. inducing or aggravating factors (such as emotion, diet, climate, drug, etc.); 8. previous history of digestive system diseases; 9. family history and allergy history; 10. combined diseases and current medication, etc.
3. Physical examination: height, weight, blood pressure, heart rate, respiratory rate, pregnancy test.
4. Disease diagnosis indicators: gastroscopy, histopathological examination, HP examination.
5. Observation time: all the above items were completed during the screening period.

### Outcome measurements

#### Primary outcome

1. Histopathological indicator Histopathological observation of chronic gastritis includes 5 histological changes and 4 grades. The grading method takes the consensus for diagnostic pathology in biopsies of chronic gastritis and epithelial neoplasms^[26]^ as a standard and is used in combination with the intuitive simulation scoring method of the New Sydney System. The 5 histological changes are: 1. HP infection; 2. chronic inflammatory reaction (infiltration of mononuclear cells); 3. activity (neutrophil infiltration); 4. atrophy (reduction of intrinsic glands); 5. intestinal metaplasia. The four grades are: 0 (none); + (mild); ++ (moderate); +++ (severe). The gastric epithelial tumor and its precursor lesions are classified into 5 grades: 1. no intraepithelial tumor; 2. uncertain intraepithelial tumors; 3. low-grade intraepithelial tumor; 4. high-grade intraepithelial tumor; 5. Cancer.
2. Evaluation method of histopathological efficacy: according to the consensus on diagnosis and treatment of CAG with TCM^[24]^, and referring to the histopathological characteristics of CAG, different variables are divided into main and secondary variables. The main variables include atrophy, intestinal metaplasia, and intraepithelial tumor, while the secondary variables include chronic inflammatory reaction, activity, and HP infection. According to the consensus for diagnostic pathology of biopsies of chronic gastritis and epithelial neoplasms^[26]^ (focusing on text description) and the intuitive simulation scoring method (focusing on picture images) of the new Sydney system, all variables are classified into “none,” “mild,” “moderate,” and “severe” 4 grades (note: grade 5, cancer in intraepithelial tumors, does not belong to the scope of this study). The 4 grades of main variables are scored as 0, 3, 6, and 9, and the 4 grades of secondary variables are scored as 0, 1, 2, and 3 respectively. Lesions often occurs in 5 parts: greater curvature of gastric antrum; lesser curvature of gastric antrum; the gastric angle; greater curvature of stomach; lesser curvature of stomach. The scores will be recorded based on the lesion in each part. The score of each variable is compared before and after treatment as the histopathological curative effect. Pathological diagnosis will report the histological changes of biopsy specimens in each site. When the multiple pathological sections in the same site is different, scores shall be given according to the most severe lesion. Table 1 show the histopathological scores.
3. Detection time: before treatment, after treatment, and during follow-up, each examination shall be conducted once for a total of 3 times.

**Table 1.** Histopathological scores.

|  |  | Grade |  |  |  | Score |
| --- | --- | --- | --- | --- | --- | --- |
|  |  | None | Mild | Moderate | Severe |  |
| Main variables | Atrophy | 0 | 3 | 6 | 9 |  |
|  | Intestinal metaplasia | 0 | 3 | 6 | 9 |  |
|  | Intraepithelial tumor | 0 | 3 | 6 | 9 |  |
| Secondary variables | Chronic inflammatory reaction | 0 | 1 | 2 | 3 |  |
|  | Activity | 0 | 1 | 2 | 3 |  |
|  | HP infection | 0 | 1 | 2 | 3 |  |
| Total score |  |  |  |  |  |  |

**Note:** The score of each variable in each part is compared before and after treatment as the histopathological curative effect.

### Secondary Outcome

#### 1. Gastroscope evaluation indicator

Gastroscopy observation: Grading standards are formulated according to the endoscopic classification and grading standard for chronic gastritis and trial opinions on treatment^[27]^. Main pathological changes: mucosal white phase, vascular exposure, plica flattening, mucosal granules, intestinal nodules. Secondary pathological changes include erosion, hemorrhage, and bile reflux. According to the endoscopic mucosal manifestations, the main and secondary pathological changes are divided into 4 grades, and the 4 grades were marked with 0, 3, 6 and 9 respectively, and the 4 grades of secondary pathological changes were scored as 0, 1, 2, and 3 respectively. Lesions often occur in the five parts (same as mentioned above), and the scores will be recorded in each part. The score of each variable is compared before and after treatment as gastroscope evaluation indicator. Table 2 show the gastroscope scores.

**Table 2.** Gastroscope scores.

|  |  | Grade |  |  |  | Score |
| --- | --- | --- | --- | --- | --- | --- |
|  |  | None | Mild | Moderate | Severe |  |
| **Main variables** | **Mucosal white phase** | 0 | 3 | 6 | 9 |  |
|  | **Vascular exposure** | 0 | 3 | 6 | 9 |  |
|  | **Plica flattening** | 0 | 3 | 6 | 9 |  |
|  | **Mucosal granules** | 0 | 3 | 6 | 9 |  |
|  | **Intestinal nodules** | 0 | 3 | 6 | 9 |  |
| **Secondary variables** | **Erosion** | 0 | 1 | 2 | 3 |  |
|  | **Hemorrhage** | 0 | 1 | 2 | 3 |  |
|  | **Bile reflux** | 0 | 1 | 2 | 3 |  |
| **Total score** |  |  |  |  |  |  |

**Note:** The score of each variable in each part is compared before and after treatment as the gastroscope evaluation indicator.

#### 2. Syndrome curative effect evaluation indicator

Nimodipine was used to evaluate the curative effect, with symptoms as the main factor, and tongue coating and pulse condition as part of the evaluation. Nimodipine calculation method: efficacy index (%) = (score before treatment-score after treatment)/score before treatment x 100%. Clinical recovery: main symptoms and signs disappear or almost disappear, with an efficacy index ≥ 95%; Obvious effect: the main symptoms and signs were significantly improved; with 70% ≤ efficacy index < 95%; Effective: main symptoms and signs improved significantly, with 30% ≤ efficacy index < 70%; Invalid: main symptoms, signs and symptoms are not significantly improved, or aggravated, with efficacy index < 30%.

#### 3. Symptom evaluation indicator

The clinical symptoms of CAG and the clinical outcomes were evaluated by using the physician report outcome scale(Clinician reported outcome, CRO)recommended by expert consensus opinion on standard quantification of spleen and stomach diseases^[28]^.

#### 4. Quality of life evaluation

In terms of quality of life, the chronic gastritis PRO (patient reported outcomes) scale, the clinical outcome evaluation scale for patients with chronic gastrointestinal diseases, the Chinese Health Status Scale and the SF-36 Health Survey Scale were used for evaluation.

#### 5. Evaluation of anxiety and depression

Hospital Anxiety and Depression Scale (HAD) was adopted to evaluate subjects who may suffer from anxiety and depression states.

#### 6. Economic evaluation

collect the medical expense data of the subjects during the study period and make cost-effectiveness analysis and cost-benefit analysis.

#### 7. Other indicators

1. Serum indicators: serum pepsinogen I, II and gastrin 17. 2. Gastric Mucosa Index: key RNA and related pathway proteins in “Inflammation-Cancer” evolution of CAG. 3. Saliva index.

#### 8. Observation time of all the above-mentioned indicators

before treatment, after treatment, and during follow-up, each examination was conducted once for a total of 3 times.

### Research evaluation Indicators

#### 1. Compliance indicators

Observation items: whether medication is taken on time and in quantity; whether other drugs are taken; whether regular follow-up visits are required; whether the observation project is completed according to the research requirements.

Evaluation method: The medication compliance of the subjects was calculated by the formula actual dosage/required dosage * 100%, and other observation contents were recorded in detail and their effects on the research results were analyzed.

Observation time: The patients were followed up once after treatment and twice during treatment.

#### 2. Combined medication index

During the study period, subjects can take drugs for the treatment of concomitant diseases. Researchers will make detailed medication records, analyze the combined medication, and explain its impact on the research results.

#### 3. Rejection and termination

Researchers will record in detail the number of cases rejected and terminated in each group and describe and record in detail the reasons for suspension, withdrawal, missed visit and rejection.

### Safety Evaluation

#### 1. Safety indicators

The subject voluntarily accepts the examination or the doctor requires the examination according to the patient’s condition.

Test items: blood analysis, urine analysis, stool routine and occult blood, electrocardiogram, liver function (ALT, AST), renal function (Cr, BUN), electrocardiogram, liver, gallbladder, spleen, pancreas and double kidney color Doppler ultrasound (the results are valid in the past month).

Researchers will record in detail the safety indicators in the research process. Abnormal safety indicators in the research process require doctors to judge its clinical significance and explain the results. If safety indicators are defined as adverse reactions or adverse events, adverse reactions and adverse events shall be recorded and reported, and corresponding countermeasures shall be taken in a timely manner.

#### 2. Safety evaluation indicators

The main indicators are both occurrence rate and detailed situation of adverse events and adverse reactions.

Detection time of all the above-mentioned indicators: before treatment, after treatment, and during follow-up, each examination was conducted once for a total of 3 times.

### Follow-up

The subjects were followed up for 24 weeks. The incidence rate of gastric mucosa high-level intraepithelial neoplasia was the endpoint outcome index. The relapse and progress of the disease were tracked for a long time to evaluate the long-term efficacy of treatment.

### Quality management of research

This study will establish standard operating procedures (SOP) for each stage in the study, such as: the recruitment process of subjects; the process of filling in eCRF; subject management; selection of treatment plan; gastroscope standard operation and sampling; gastric mucosa specimen processing and transportation; handling and transporting serum samples; handling and transporting saliva samples. All SOPs will be entered into the electronic database. In addition, this study will carry out regular maintenance and testing of the equipment needed in the study to avoid laboratory errors.

### Data management

Researchers need to ensure the authenticity and integrity of the data in the case report form and treat the inevitable missing data according to the missing values in statistics.

### Statistical analysis

#### 1. Descriptive statistics

SPSS 17.0 software was used for calculations. Counting data are expressed by frequency and composition ratio. Measurement data conform to the mean and standard deviation of normal distribution, while measurement data that do not conform to normal distribution are expressed by median or mode.

#### 2. Difference test

SPSS 17.0 software was used for calculations, and chi-square was used for comparison between counting data groups. Covariance analysis/repeated measurement analysis of variance is used when the normal distribution is compared between measurement data groups and the variance is uniform. Paired rank sum test is used for non-normal distribution or uneven variance.

## Discussion

To date, the treatment of CAG still revolves around the eradication of HP, antacids, prokinetics and mucosal-protective agents^[29]^. However, HP has developed resistance to several antibiotics^[30]^, which means that HP eradication may be ineffective in some HP positive patients. TCM treatment can provide a powerful and safe therapeutic approach for the treatment of CAG.

According to the Consensus on TCM Diagnosis and Treatment of Chronic Gastritis (2017)^[19]^, FSCHABM is appropriate for the entire treatment process of CAG. In clinical practice, TCM practitioners will flexibly change Chinese herbs and prescriptions according to the patient’s symptoms and signs, but the method of treatment always revolves around FSCHABM.

Regarding CAG’s existing RCT report, however, CAG has a short treatment course and is at high risk of selection and performance bias due to a lack of reporting of information regarding allocation concealment and blindness, especially for TCM research. RCT strictly requires random grouping and to give medicines uniformly, which contradicts the concept of holism and treatment based on syndrome differentiation, and fails to give full play to the curative effect of TCM. To pander to the basic theory of TCM, and pursue high-quality clinical trials, a new model of research is required for truly reflecting the efficacy of TCM. Undoubtably, the real-world study is consistent with the clinical practice of TCM, and is also suitable for chronic disease research. As a chronically developed disease, CAG urgently needs this research model to explore the development and efficacy of the disease treatment. However, the real-world study of CAG has not been reported up to now. The treatment of FSCHABM after repeated clinical practices shows that it is always effective in improving the symptoms of CAG patients. However, the occurrence and development of CAG, the curative effect and therapeutic mechanism of FSCHABM are unclear. Therefore, the purpose of this protocol is to evaluate the clinical efficacy and safety of FSCHABM in the treatment of CAG. We conducted a clinical research and comprehensive evaluation of TCM’s intervention for CAG and explored the molecular mechanism of FSCHABM in the treatment of CAG. Moreover, this protocol is designed to provide an excellent clinical study protocol for clinical researches of CAG or other chronic diseases, which is a more realistic display of the development and curative effect of TCM, and provides a reference for the multi-center, randomized, controlled trials.

### Trial status

The protocol version number is 2.0. Trial recruitment was started on March 30, 2018, and the trial has enrolled 458 patients at the time of manuscript submission. Recruitment is expected to be completed in March 2028.

### Ethics and dissemination

Study findings will be shared with participants, healthcare providers, and policymakers through research reports, conference presentations, and the Internet. The results will also be disseminated through publication in peer reviewed journals.

## Abbreviations

CAG: Chronic atrophic gastritis
HP: *Helicobacter pylori*
FSCHABM: the fortifying the spleen, clearing heat, activating blood method
TCM: traditional Chinese medicine
RCT: randomized controlled trial
PRO: patient reported outcomes
HAD: Hospital Anxiety and Depression Scale
SOP: standard operating procedures

## Authors’ contributions

Zhihua Zheng and YI Wen designed the study and drafted the manuscript; Peiwu Li and Jinglin Pan provided methodological advice and critically revised the manuscript; Yanhua Yan, Zhiheng Xu, Kecao Nie, Xu Chen, Fengbin Liu were involved with recruitment and follow-up; All authors have read and approved the final version of the manuscript.

## Acknowledgements

We wish to thank doctor Fengbin Liu from the first Affiliated Hospital of Guangzhou University of Chinese Medicine for providing experimental guidance.

---

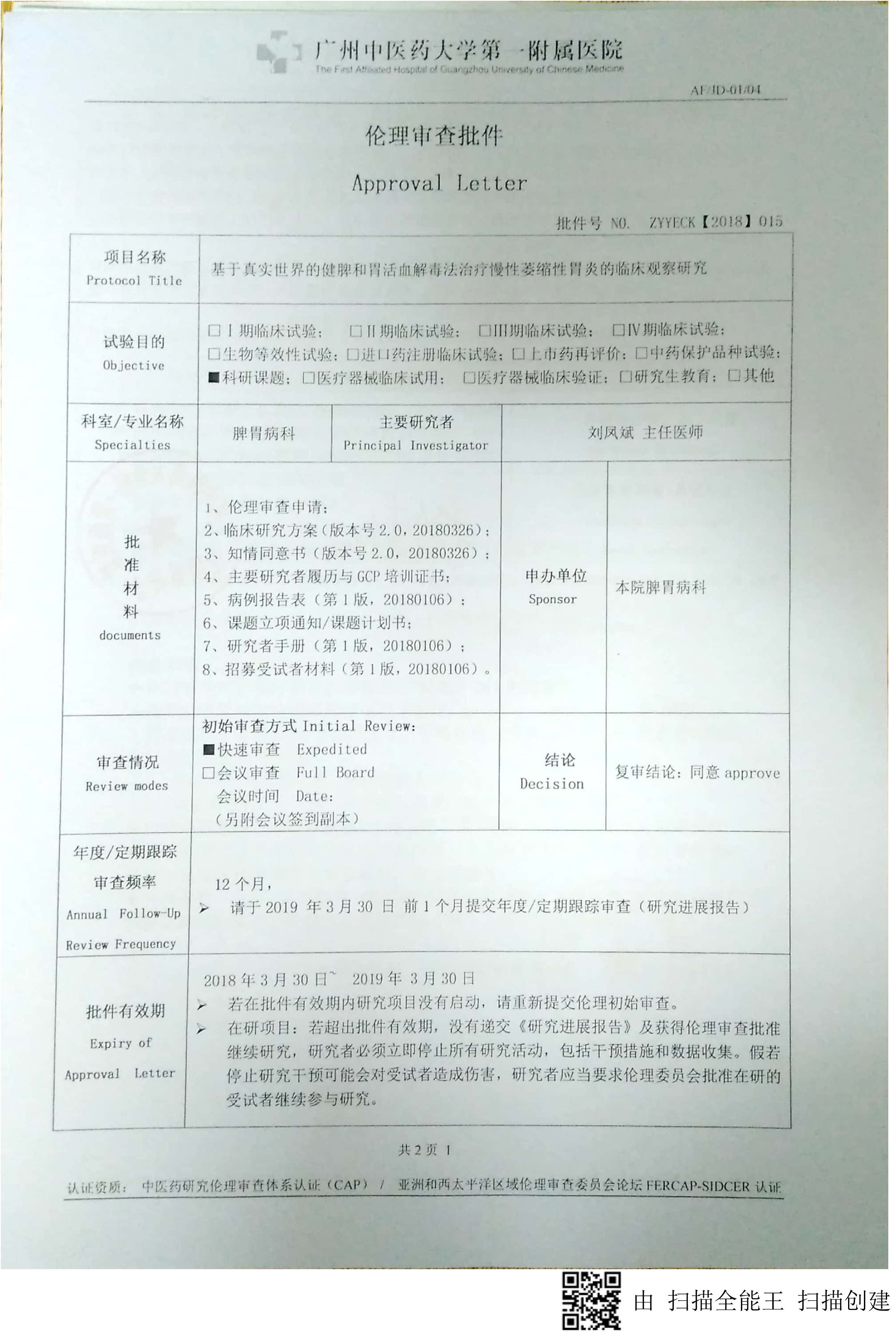
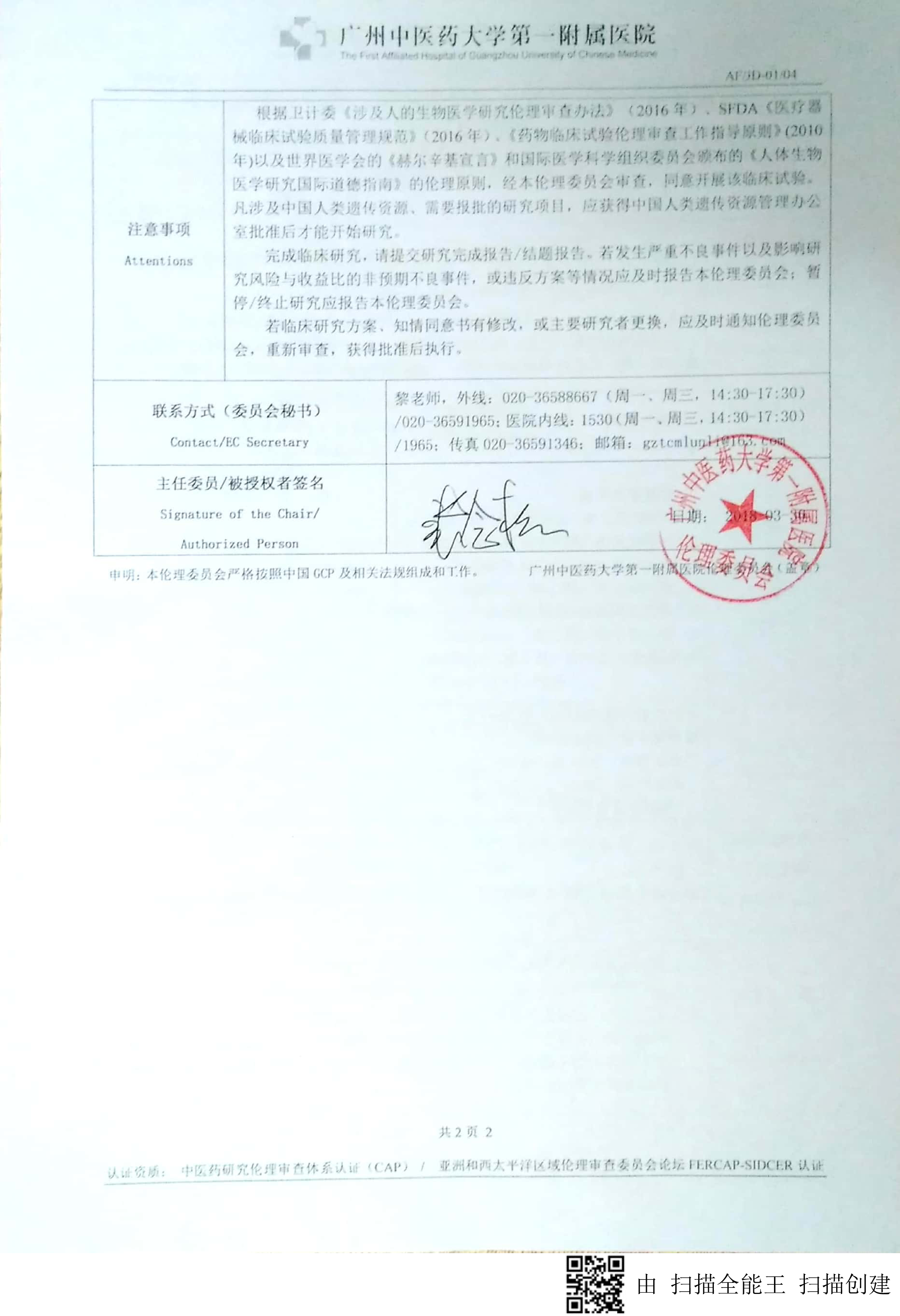
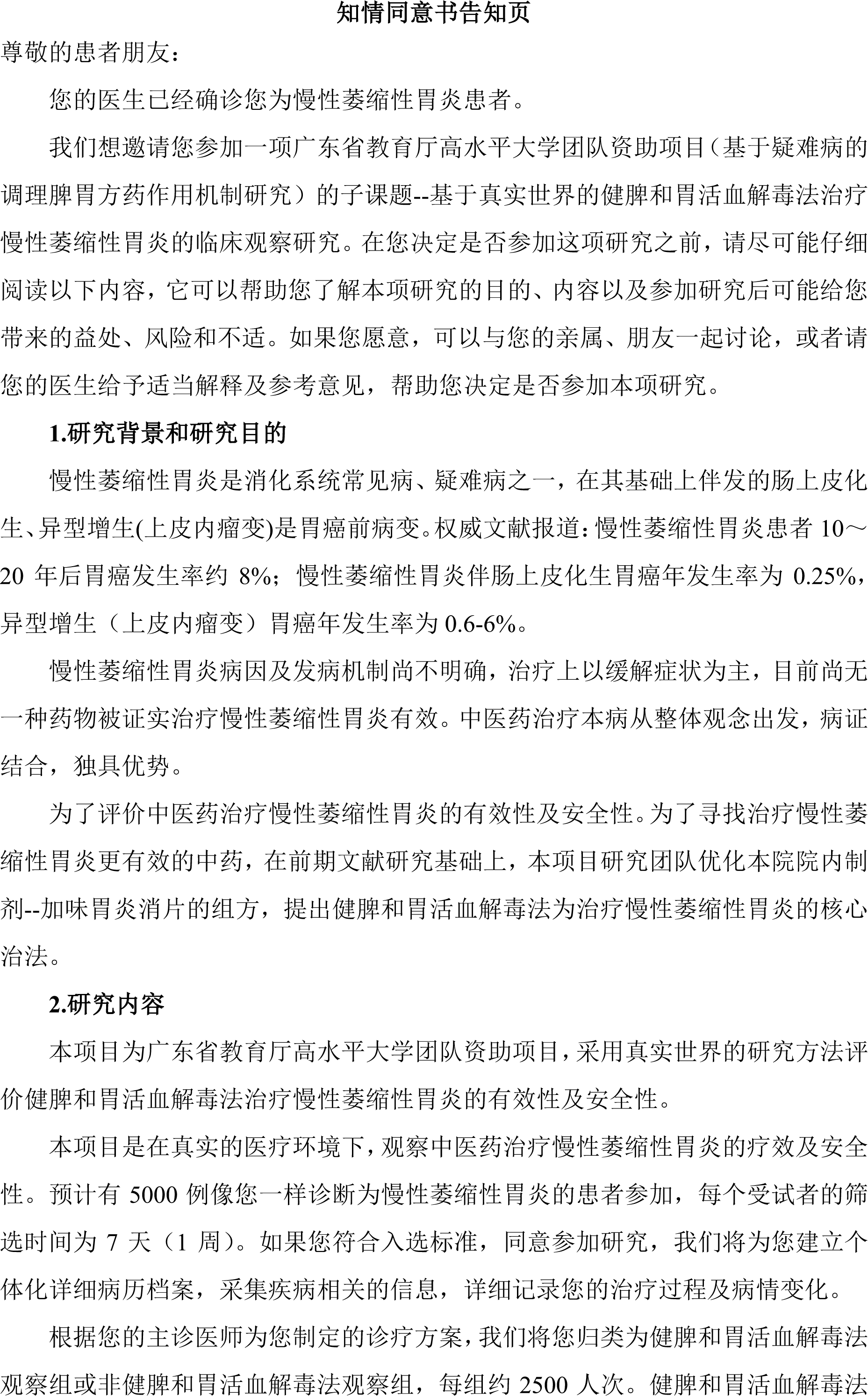
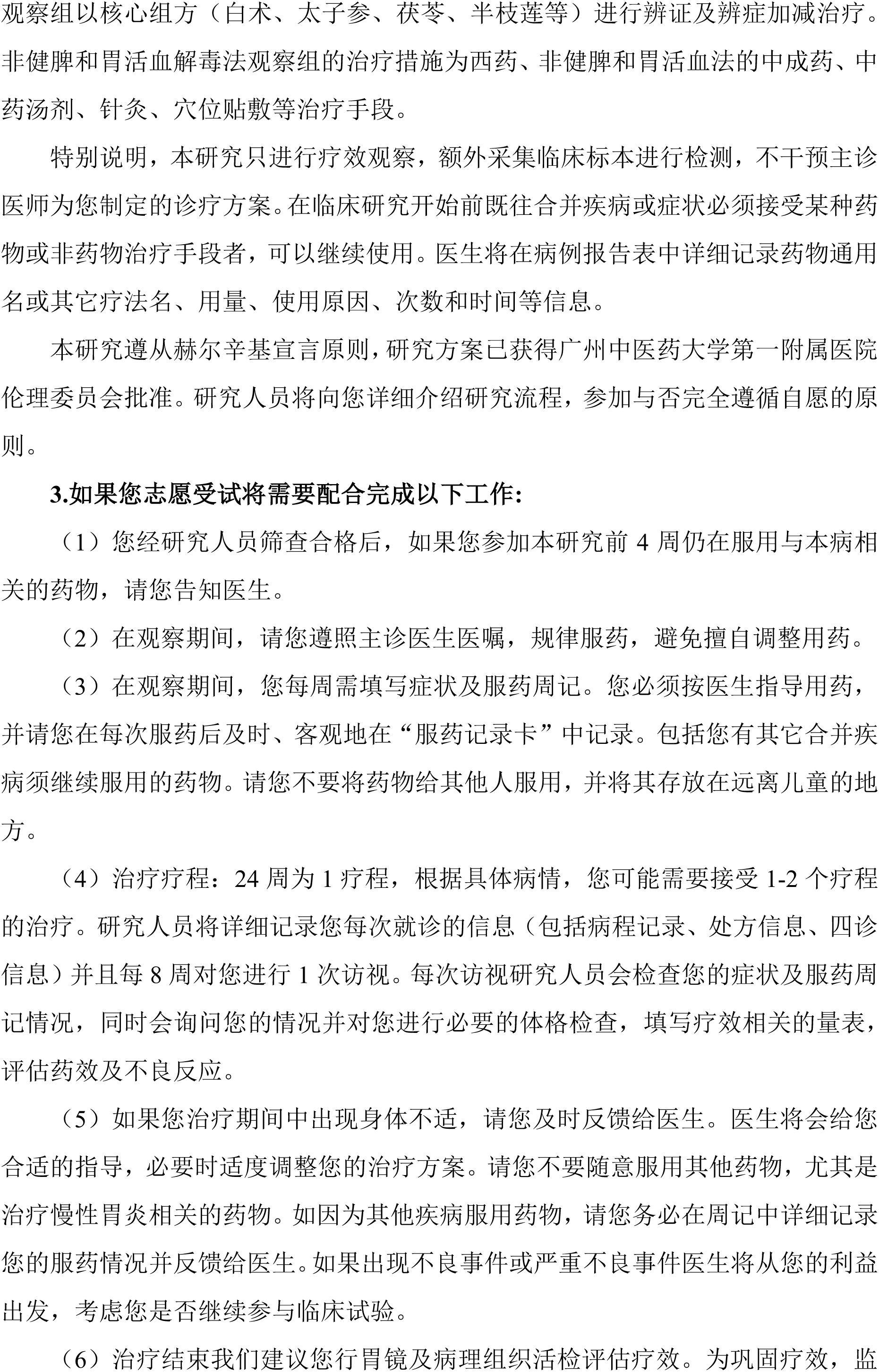
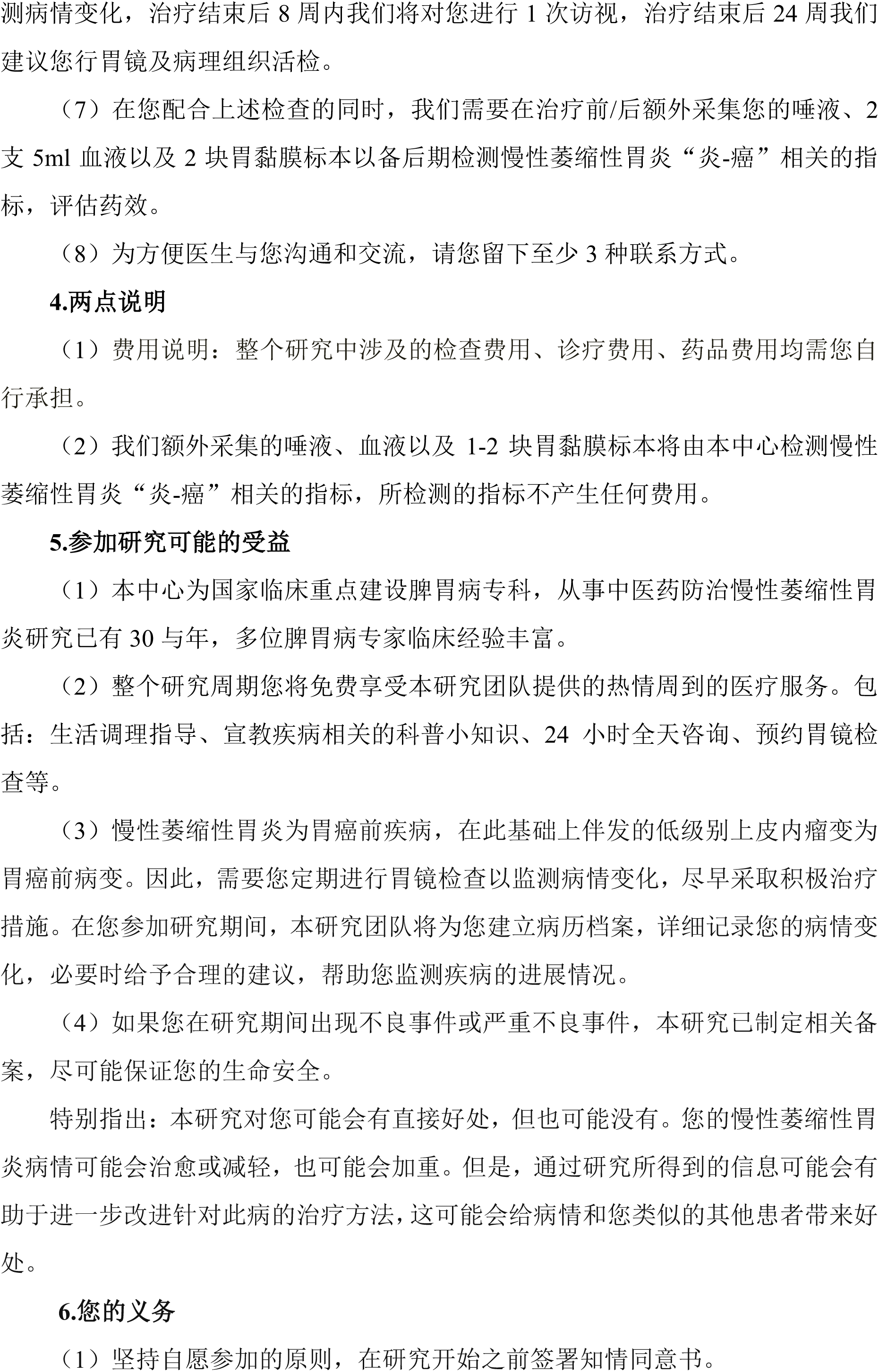
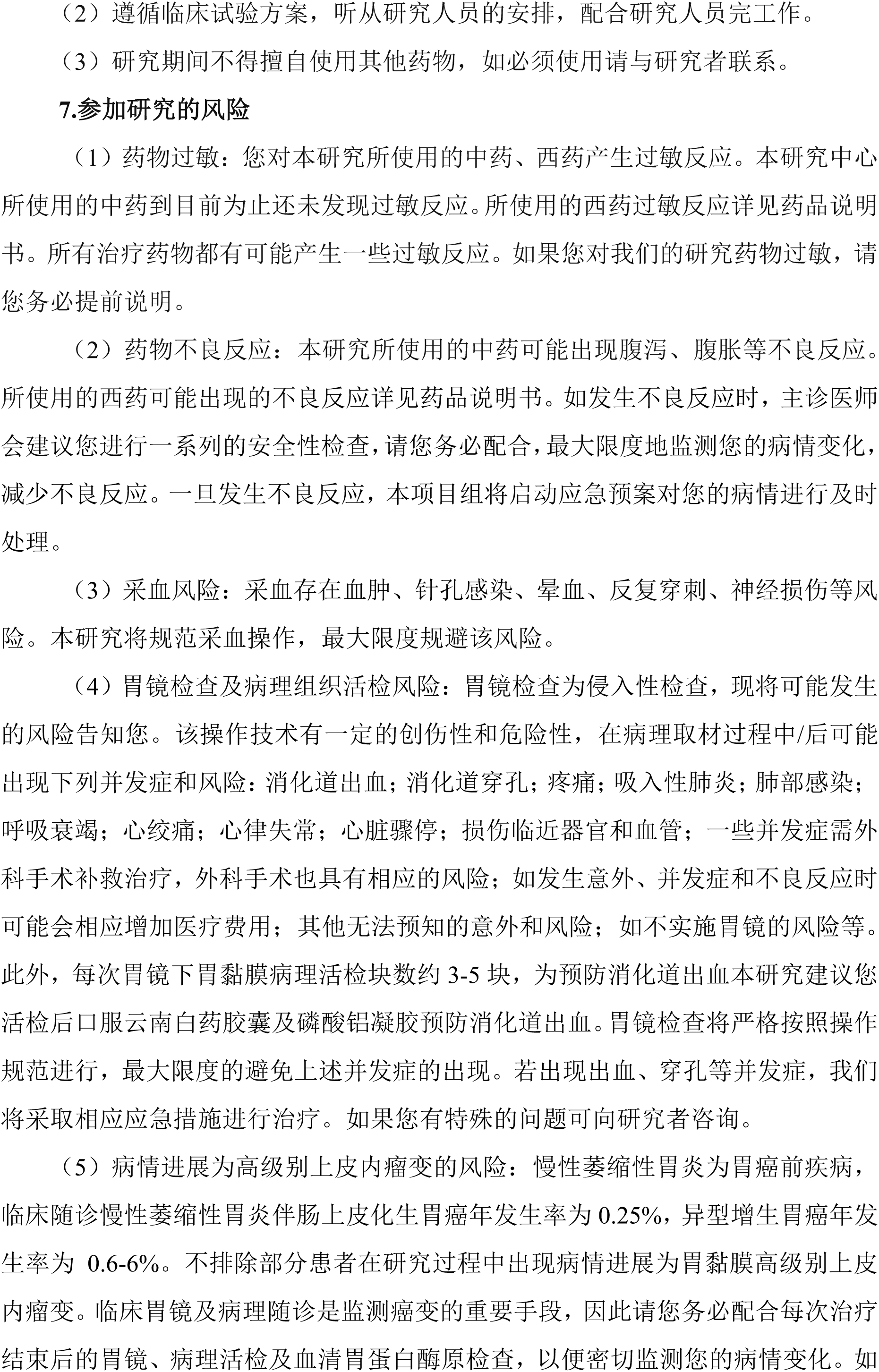
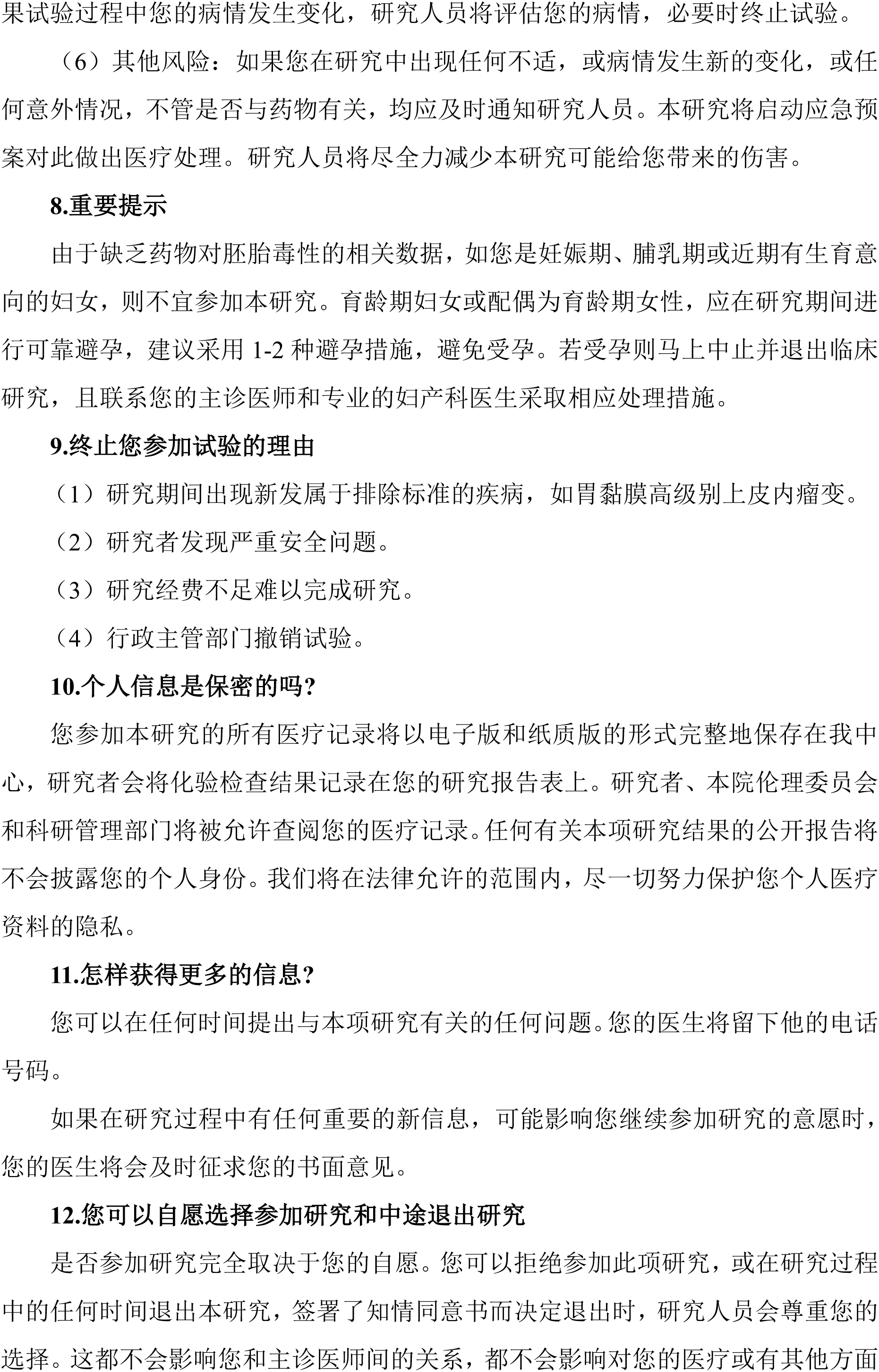
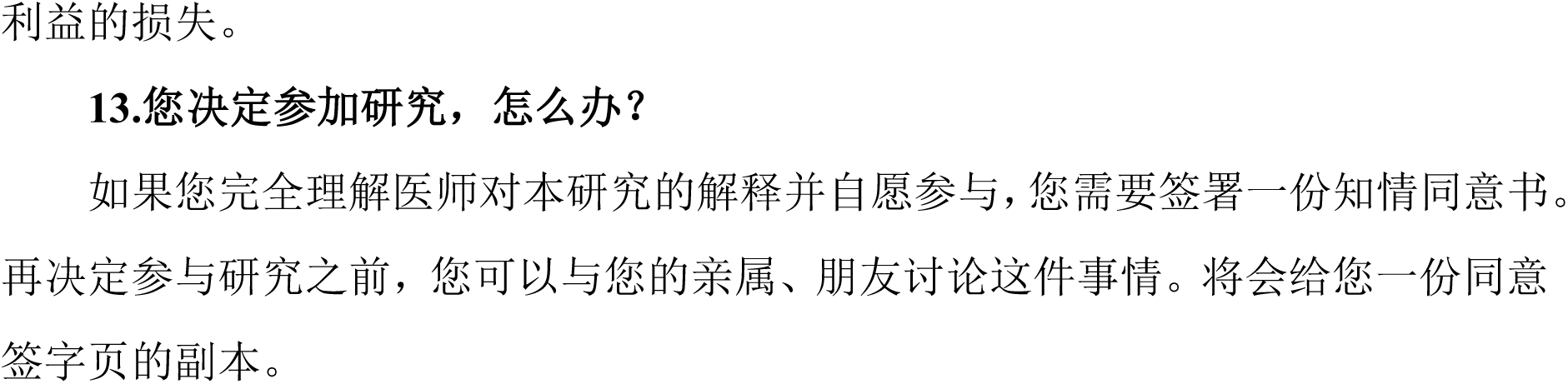
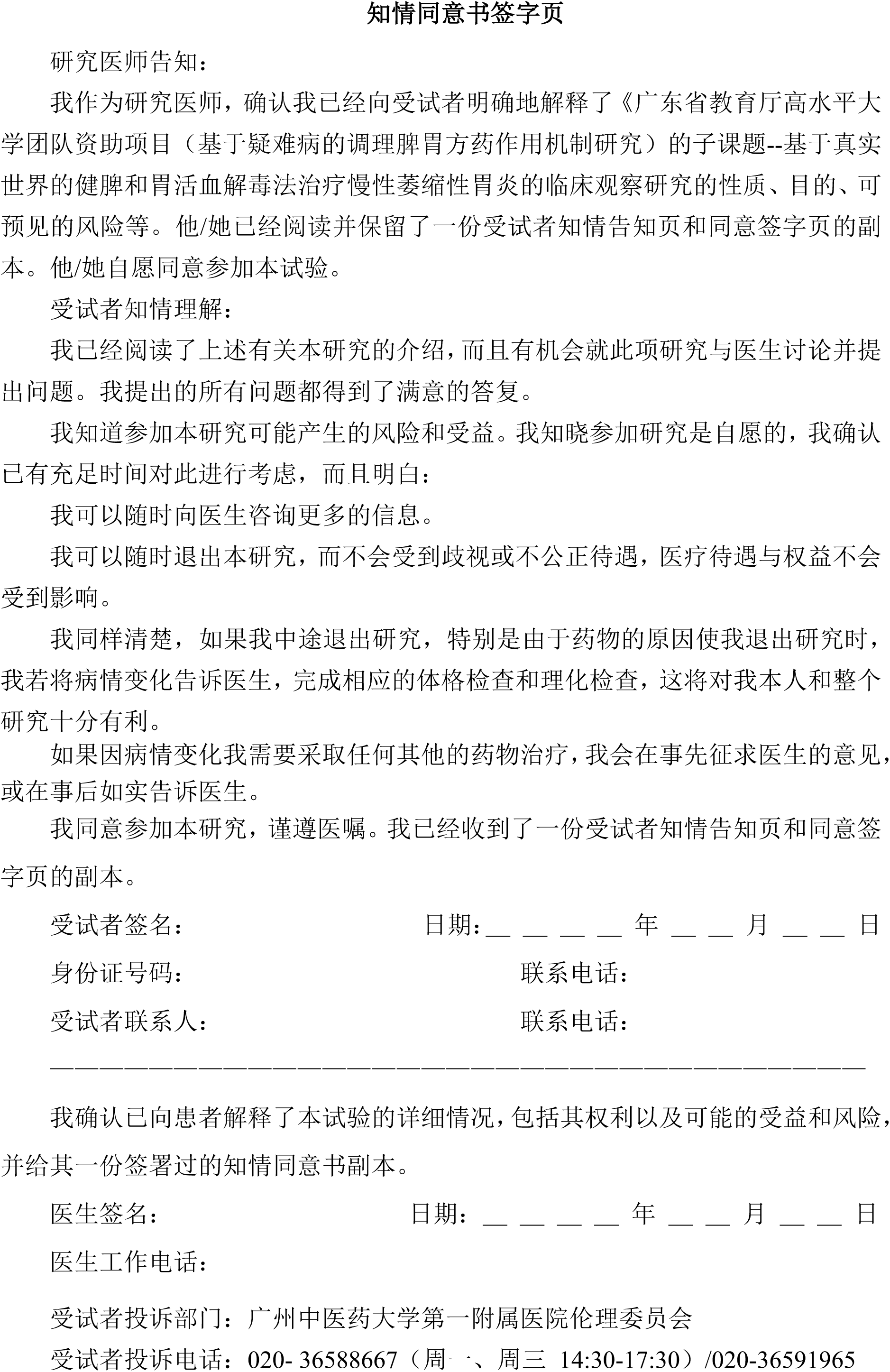

